# Closed-loop control of k-space sampling via physiologic feedback for cine MRI

**DOI:** 10.1101/2020.06.22.20137638

**Authors:** Francisco Contijoch, Yuchi Han, Srikant Kamesh Iyer, Peter Kellman, Gene Gualtieri, Mark A. Elliott, Sebastian Berisha, Joseph H. Gorman, Robert C. Gorman, James J. Pilla, Walter R. Witschey

## Abstract

**Background:** Segmented cine cardiac MRI combines data from multiple heartbeats to achieve high spatiotemporal resolution cardiac images, yet predefined k-space segmentation trajectories can lead to suboptimal k-space sampling. In this work, we developed and evaluated an autonomous and closed-loop control system for radial k-space sampling to increase sampling uniformity.

**Methods:** The closed-loop system autonomously selects radial k-space sampling trajectory during live segmented cine MRI and attempts to optimize angular sampling uniformity by selecting views in regions of k-space that were not previously well-sampled. Sampling uniformity and robustness to arrhythmias was assessed using ECG data acquired from 10 normal subjects in an MRI scanner. The approach was then implemented with a fast gradient echo sequence on a whole-body clinical MRI scanner and imaging was performed in 4 healthy volunteers. The closed-loop k-space trajectory was compared to random, uniformly distributed and golden angle view trajectories via measurement of k-space uniformity and the point spread function. Lastly, an arrhythmic dataset was used to evaluate a potential application of the approach.

**Results:** The autonomous trajectory increased k-space sampling uniformity by 13±7%, main lobe point spread function (PSF) signal intensity by 14±6%, and reduced ringing relative to golden angle sampling. When implemented, the autonomous pulse sequence prescribed radial view angles faster than the scan TR (0.98 ± 0.02 ms, maximum = 1.38 ms) and increased k-space sampling mean uniformity by 5±12%, decreased uniformity variability by 45±14%, and increased PSF signal ratio by 5±5% relative to golden angle sampling.

**Conclusion:** The closed-loop approach enables near-uniform radial sampling in a segmented acquisition approach which was higher than predetermined golden-angle radial sampling. This can be utilized to increase the sampling or decrease the temporal footprint of an acquisition and the closed-loop framework has the potential to be applied to patients with complex heart rhythms.

## Background

Cine MRI captures the motion of the heart by acquiring images at frame rates faster than the motion occurs. A straightforward approach to this problem is to collect the image at a frame rate much higher than the heart rate [1,2], yet practical limitations of physiology, hardware and patient safety limit how quickly image data can be obtained and spatial or temporal fidelity may be compromised. The lowest acceptable frame rate to visualize a heart beating at 60 beats-per-minute is about 20 frames-per-second and higher frame rates are required for higher heart rates [3]. To improve fidelity, cine MRI is conventionally performed using segmented sampling techniques where periodic motion enables a subset of k-space data to be collected [4–6].

Segmented MRI can be performed using different k-space trajectories including Cartesian and non-Cartesian patterns. However, Cartesian trajectories are known to be sensitive to arrhythmias and data from irregular beats must be reacquired otherwise images will have inconsistent spatial information and artifacts. While arrhythmia rejection algorithms have been developed, rejecting data if a heartbeat is too long or short and reacquiring it in the subsequent beat can lead to breathholds that are too long for patients. Alternatively, non-Cartesian radial [7–10] and spiral trajectories [11,12] have the advantageous properties of local k-space uniformity, meaning that adjacent lines of data can be distributed uniformly in k-space, and variable density sampling. In particular, golden angle acquisitions [13] can address the problem of arrhythmias by using an adaptive temporal footprint [14,15]. However, a limitation of this approach is that only a contiguous set of golden angle views will have uniform k-space sampling properties. As a result, segmented golden angle trajectories are suboptimal since views collected from adjacent heartbeats will not have near uniform k-space sampling [14,16,17].

To address this limitation, we developed a radial trajectory that adapts in response to physiologic changes in the patient being scanned and uses same-scan data to optimize the sampling trajectory on-the-fly using a closed-loop. Sampling uniformity and point spread function signal properties were evaluated using ECG data from subjects with normal rhythm and in one patient with an arrhythmia. We implemented the autonomous control system on a whole-body clinical MRI scanner. We show feasibility of this approach in 4 healthy human volunteers and demonstrate initial proof of utility in patients with complex rhythms. Images and sampling properties were compared to a conventional segmented golden angle cine MRI.

## Methods

### Closed-loop radial k-space sampling

We developed a closed-loop radial acquisition in which the k-space trajectory to be acquired is calculated dynamically throughout the scan. The data to acquire is determined according to the segmented data which was previously acquired with the goal of minimizing angular gaps in k-space and reducing unequal angular sampling density. All acquired k-space locations are time-stamped and a simple cross-correlation of the ECG signal is used to identify prior periods of similar cardiac phase. The advantage of ECG-matching by cross-correlation is that it has real-time performance and can compute cross-correlation results in less than 1 msec - an essential requirement for fast scans such as segmented cine MRI which requires very rapid repetition times.

An overview of the closed-loop radial k-space sampling acquisition is shown in **Figure 1** and more details are included in **Supplemental Figure 1**. Prior periods of similar ECG signal are identified as local maxima of the cross-correlation of the historical signal with the most recent ECG signal. In conventional segmented k-space trajectories, the total number of projections *N*_*θ*_ is obtained by sampling a subset of radial views (segments *N*_*s*_) during each heartbeat (shots *N*_*q*_). **Figure 1** shows how segments and shots are defined for a closed-loop acquisition. In our closed-loop approach the definition of *N*_*θ*_ is different from a traditional segmented trajectory, for which *N*_*θ*_ = *N*_*s*_*N*_*Q*_, as closed-loop sampling can only use half the views of the current shot since the other half has not occurred yet. For the four heartbeat segmented cine example shown in **Figure 1** there are 3 shots with 4 segments (i.e. radial views), but the fourth shot (yellow) has only 2 views.

**Fig 1.**
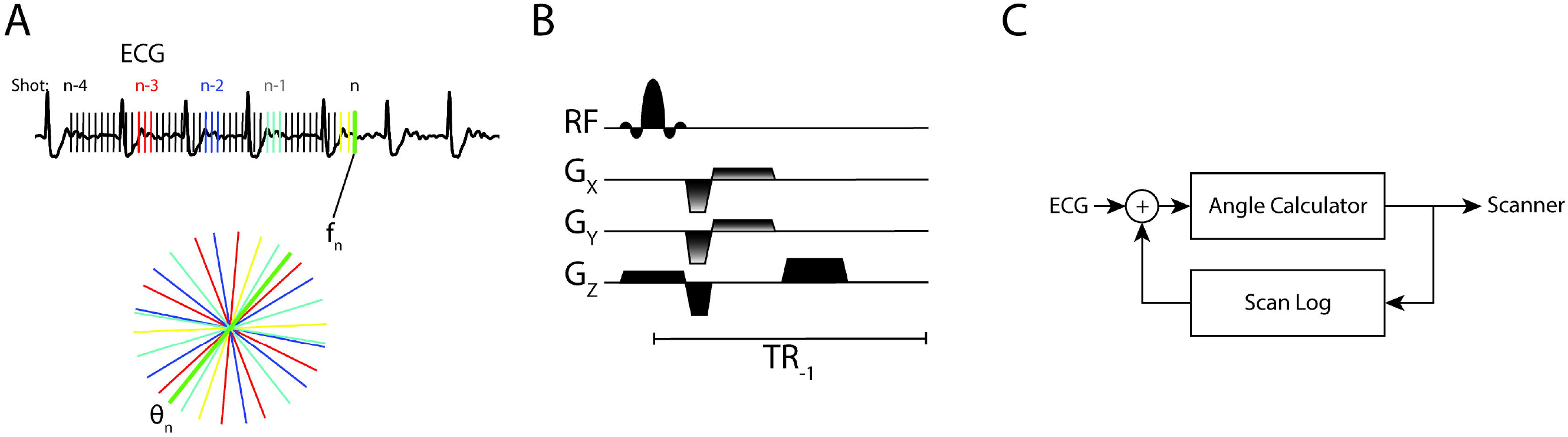
Segmented acquisition using an autonomous closed-loop system **A** During the acquisition, the closed-loop system identifies prior, similar phases of the cardiac cycle using cross-correlation of the recorded ECG signal. For frame f_n_, radial k-space lines from prior phases (red, blue, and teal) as well as the most recent views (yellow) are aggregated and the newest projection θ_n_ (green) is defined to bisect the largest angle, thus closing the gap in k-space. **B** The calculated angle θ_n_ is provided to the bSSFP sequence in real-time such that the Gx and Gy gradients are updated. **C** The closed loop nature of the physiologic signal (ECG), the angle calculator, scanner, and log of prior scan data.

From the list of all previously acquired view angles, the subset corresponding to the periods associated with local maxima in the cross-correlated ECG signal are collated. The new radial angle *θ*_*i*_ (green radial line in **Figure 1A**) is chosen so that it bisects the largest angular gap Δ*θ*_*i*_. As a result, the closed-loop acquisition closes the largest gap in k-space and improves angular sampling uniformity.

### Implementation and interface of closed-loop radial scheme with MRI system

To demonstrate the feasibility of autonomous radial imaging, we developed a software platform for closed-loop radial imaging and interfaced it to a whole-body clinical MRI scanner. As outlined in **Supplemental Figure 2**, the system consisted of four systems for real-time communication and feedback: (1) the physiologic monitor, (2) adaptive measurement controller, (3) pulse sequence, and (4) digital signal processors (DSPs).

ECG signals were received from the patient using a MR-compatible 4-lead system (In vivo Gainesville, FL USA). The ECG signal was transmitted wirelessly to the physiologic monitor. Analog signals were digitized, logged and transmitted via TCP/IP in software (LabView, National Instruments, Austin, TX USA) to the adaptive measurement controlled, which in turn determined a new view angle.

During the closed-loop period, the adaptive measurement controller and pulse sequence respond to ECG feedback. The adaptive measurement controller performs 3 steps sequentially: a) transmits an updated k-space trajectory to the pulse sequence; b) reads and stores in memory new ECG data; c) analyzes ECG data and computes a new k-space trajectory. The pulse sequence receives the new trajectory from the adaptive measurement controller, transmits the instructions to the DSPs and enters the standby period until the DSPs have executed a wake-up instruction. After wake-up, the pulse sequence waits for a new trajectory from the adaptive measurement controller and repeats.

To evaluate real-time timing performance, the calculation times for the adaptive measurement controller were measured for the initialization and active periods of the software. Average, standard deviation, and maximum calculation times were calculated. Timing information was calculated during scanning of 4 subjects. For each subject, 4 scans were performed according to the segmentation strategies in **Table 1**. The average calculating time for the initialization period was 0.792 ± 0.028 ms and for active mode was 0.982 ± 0.017. The maximum time to update was 1.11 ms during the initialization period and 1.375 msec during the active period was. All maximum update times were faster than the repetition time of the sequence.

**Table 1:** Performance of closed-loop radial imaging sampling for 10 subjects with recorded ECG for different k-space segmentation sampling schemes

| $N_\theta$ | $N_q$ | $N_s$ | Uniformity (%)<sup>s</sup> | | | PSF Signal Ratio | | |
| --- | --- | --- | --- | --- | --- | --- | --- | --- |
|  |  |  | ARKS | Golden | Random | ARKS | Golden | Random |
| 27 | 1 | 54 | 94.6±1.0 | 82.9±0.0 | 50.9±0.1 | 99.7±0.4 | 89.1±0.0 | 86.5±0.4 |
|  | 2 | 18 | 71.5±0.6 | 66.9±1.5 | 50.9±0.1 | 93.4±0.3 | 85.6±0.7 | 86.7±0.6 |
|  | 5 | 6 | 69.0±1.1 | 56.5±1.8 | 50.9±0.1 | 92.0±0.5 | 82.1±0.8 | 86.5±0.3 |
| 45 | 1 | 90 | 94.4±1.1 | 83.8±0.0 | 50.6±0.2 | 99.2±0.5 | 85.7±0.0 | 79.0±0.4 |
|  | 2 | 30 | 70.4±0.7 | 66.1±3.4 | 50.6±0.1 | 88.8±0.5 | 82.2±0.9 | 78.7±0.5 |
|  | 5 | 10 | 68.0±1.1 | 56.3±3.5 | 50.5±0.1 | 86.6±0.8 | 77.1±1.5 | 78.9±0.4 |
|  | 8 | 6 | 66.5±1.3 | 54.3±2.4 | 50.6±0.1 | 85.8±1.1 | 76.2±1.0 | 78.7±0.6 |
| 75 | 1 | 150 | 94.4±0.8 | 82.0±0.0 | 50.3±0.1 | 99.0±0.6 | 80.9±0.0 | 75.0±0.7 |
|  | 2 | 50 | 69.5±0.6 | 64.7±4.3 | 50.4±0.2 | 86.7±1.0 | 78.7±0.6 | 74.8±0.4 |
|  | 8 | 10 | 65.8±1.0 | 53.5±4.3 | 50.3±0.1 | 83.2±0.8 | 72.1±1.5 | 74.9±0.4 |

<sup>s</sup>Uniformity (Eq. 12) compares the performance of the radial acquisition to the ideal distribution (Eq. 9). $N_\theta$ = number of radial views, $N_q$ = number of shots, $N_s$ = number of segments/shot, ARKS = autonomous radial k-space sampling, GA = golden angle, PSF = point spread function.

### Simulation-based assessment of closed-loop k-space trajectory performance

We performed simulations using recorded ECG signals to investigate the distribution of view angles that would be assigned under a closed-loop radial trajectory. This simulation did not include k-space or image space data but was used to evaluate the distribution of view angles for different ECGs with respect to angular sampling density (uniformity) and their point spread function (PSF). For this simulation, 3-lead, chest ECG data was collected from 10 normal subjects at a 400 Hz sampling rate and resampled to the MRI scanner repetition time (TR). The duration of the ECG recordings was 75.7 ± 23.7 sec, corresponding to 2.7 ± 0.85 x10^4^ views at TR = 2.8 ms.

After an initial training period were golden angle radial sampling *θ* = 111.25° occurred, ECG signals were cross-correlated, and view angles were determined for the segmentation schemes shown in **Table 1**. These results were compared to segmented golden angle and random radial approaches. Golden angle views continuously incremented at the golden angle following the initialization period. Random view angles were chosen from the interval 0 to 180° with uniform (flat) probability distribution for selection. In this study, all subjects and patients gave informed consent prior to participating in the study, approved by the Institutional Review Board of the University of Pennsylvania.

A second set of closed-loop radial simulations were performed using ECG and previously collected cardiac cine data from one patient with an arrhythmia. Image data was acquired on a 1.5 T whole-body MRI system (Avanto; Siemens Healthcare; Erlangen, Germany) equipped with a 40 mT/m gradient coil and a 32 channel RF receiver array (16 anterior and 16 posterior elements). Cardiac gating was obtained with a 3-lead wireless ECG system. Time-stamps were communicated using TCP/IP from the pulse sequence to the ECG log file to synchronize imaging and ECG data. Left ventricular, short-axis, real-time data was obtained using a golden angle radial trajectory and image parameters, flip angle = 70°, TE = 1.4 ms, TR = 2.8 ms, number of frequency encoded points = 128, field-of-view = 340 mm x 340 mm, slice thickness = 8 mm, bandwidth = 1140 Hz/pixel. 10-40 seconds of continuous golden angle radial data was collected, resulting in 6000-20000 golden angle radial projections. K-space signal data was reconstructed offline using adaptive coil synthesis [18] and non-Cartesian SENSE algorithm [19] in open-source software [20] on a Linux workstation as previously detailed [21]. The reconstructed image frame rate was 300 frames per second and exposure time (temporal footprint) was 95 ms (= 34 projections per frame). To remove residual radial streak artifacts, a median filter was applied with a width of 30 frames. The final 128 x 128 images were interpolated to 512 x 512 to generate the simulation spin density *ρ* (***x***). Simulated k-space data was then generated and sampled using random, golden angle or closed-loop acquisitions. Images were reconstructed as for the in vivo data in the following sections.

### In vivo evaluation of closed-loop MRI imaging and image reconstruction

Imaging was performed on a 1.5 T whole-body MRI scanner (Avanto, Siemens Healthcare, Erlangen, Germany) with a 16-channel RF receiver array. 4 healthy volunteers subjects participated in this study. To visualize cardiac contraction, a mid-ventricular short-axis slice was imaged. Subjects were imaged using a spoiled gradient echo sequence with the following parameters: TE/TR=4.1/8.2 ms, FOV = 320×320 mm^2^, bandwidth = 240 Hz and flip angle = 12°. The MRI sequence sent a request to the adaptive controller for the next radial angle to acquire 2 ms prior to the end of the current data acquisition (every TR). This 2 ms window allowed for reply from the adaptive controller as well as preparation of the RF gradients by the scanner given the prescribed angle. Acquisitions were performed with various combinations of radial views (segments) per heartbeat (shots) as shown in the **Table 2**.

**Table 2:** Mean and variability of k-space uniformity for golden-angle and autonomous scanning

| N<sub>q</sub> | N<sub>s</sub> | Mean Uniformity |  | Uniformity Variability |  | PSF Signal Ratio |  |
| --- | --- | --- | --- | --- | --- | --- | --- |
|  |  | GA | ARKS | GA | ARKS | GA | ARKS |
| 5 | 6 | 56.4 ± 5.9 | 57.7 ± 0.4 | 8.0 ± 2.4 | 5.4 ± 0.2* | 80.5 ± 2.8 | 82.3 ± 0.9 |
| 5 | 10 | 53.0 ± 7.1 | 62.7 ± 1.6* | 9.7 ± 3.3 | 4.1 ± 0.1* | 72.3 ± 5.0 | 79.8 ± 0.8* |
| 8 | 6 | 58.3 ± 4.5 | 52.8 ± 1.4* | 6.5 ± 1.1 | 4.4 ± 0.3* | 75.3 ± 2.5 | 77.2 ± 0.5 |
| 8 | 10 | 56.5 ± 6.5 | 61.4 ± 1.1 | 7.2 ± 3.1 | 3.2 ± 0.1 | 68.9 ± 3.8 | 73.3 ± 1.4* |

\*indicates significant ( $p < 0.05$ ) difference between GA and adaptive approach. N<sub>q</sub> = number of shots, N<sub>s</sub> = number of segments/shot, ARKS = autonomous radial k-space sampling, GA = golden angle, PSF = point spread function.

Image reconstruction was performed using a previously-described iterative reconstruction with spatial and temporal variation constraints which also incorporates parallel imaging [22,23]. Weights were empirically tuned on a test dataset to provide high visual quality, while achieving rapid convergence of the cost function. The iterative approach was stopped when the convergence criterion was reached:

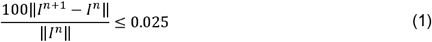

### Sampling Performance Metric: Point spread function analysis

The number of projections *N*_*θ*_ required to fulfill Nyquist sampling and prevent aliasing of the Fourier signal is [24,25]

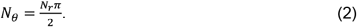

where *N*_*r*_ is the number of samples *k*_*r*_ per projection. More generally, the point spread function (PSF) describes degradation of MR image by the k-space sampling trajectory. The PSF for radial k-space sampling trajectories with uniform sampling density is minimally degraded inside a circular field-of-view [26]

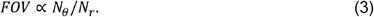

If *N*_*θ*_ ≪ *N*_*r*_, then the circular FOV is reduced and signal contamination occurs.

In general, Eq. (2) and (3) are correct for a radial sampling distribution in which the view angles are separated by a single angle Δ*θ*. However, while other radial sampling trajectories may not satisfy the same Nyquist sampling criteria given by Eq. (2) [26], it is still possible to understand the aliasing properties of the image from the PSF of the sampling trajectory such as for closed-loop sampling. An additional complicating factor is that each image frame does not have the same distribution of Δ*θ* so it is not possible to understand all the aliasing properties of closed-loop radial sampling from a single image frame.

To better understand potential image space artifacts caused by closed-loop radial sampling and segmented golden angle imaging, we estimated the PSF for each image frame individually and combined these individual results to present a single average PSF. We observed that these average results showed circular symmetry on account of the large number of image frames that were involved. The circularly symmetric PSF could thus be presented as a single 1D projection through the center of the 2D PSF. Revolving the 1D circularly symmetric PSF around the coordinate [*k*_*x*_ = 0, *k*_*y*_ = 0] by 180° would recover the original 2D PSF. The PSF signal intensity ratio was estimated from the 1D circularly symmetric PSF as the sum of signal s(x) in the main lobe (-*τ* to *τ*) divided by the total signal in the image.

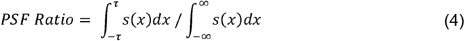

### Sampling Performance Metric: Quantitative Analysis of View Angle Distribution

We analyzed the distribution of view angles for different k-space sampling approaches using probability and cumulative distribution functions as illustrated by **Supplementary Figure 3**.

In general, it should be noted that radial k-space trajectories do not uniformly sample k-space, since the center of k-space has higher sampling density than the periphery. The ideal distribution for radial k-space sampling is only ideal with respect to the *θ* parameter.

The deviation from the ideal distribution is given by the cumulative distribution function

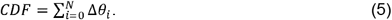

The approximate and ideal CDFs were then compared

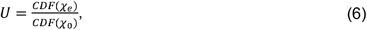

where *CDF* (χ_*e*_)is the cumulative distribution of the comparison trajectory and *CDF* (χ_0_)is of the ideal trajectory. *U* is the uniformity of the sampling distribution and quantitatively compares any view angle distribution to the ideal distribution. Analytical expression for Eqs. (5-6) are not available for closed-loop radial imaging since it depends on the ECG, algorithm and acquisition parameters. Nevertheless, it is possible to estimate an approximate distribution by repeated random sampling of ECG data using known scan parameters.

### Statistics

Continuous, quantitative measures such as angular uniformity and PSF ratio are reported as mean values with standard deviations. Significant differences in angular uniformity and PSF signal ratio between the proposed closed-loop system and golden angle and random sampling were estimated using paired, two-tailed t-tests at a P < 0.05 level of significance (Matlab, the MathWorks, Natick, MA).

## Results

### Closed-loop, segmented cine MRI acquisition

Cardiac cine MRI data was acquired on 4 healthy subjects using the autonomous sampling trajectory at 1.5 T. **Figure 2** shows results from a segmented (four-shot) acquisition during a breathhold in a normal subject. The adaptive sampling technique showed sampling uniformity across the cardiac phase. Furthermore, the image quality was good and showed that end-systolic and end-diastolic periods were well-resolved with good ventricular-blood contrast.

**Fig 2:**
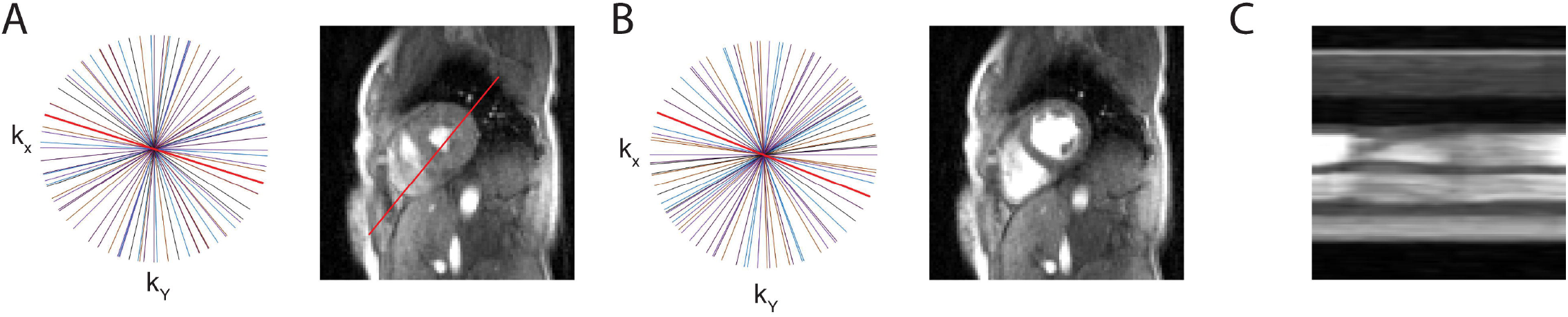
Closed-loop radial sampling of scanning in a healthy volunteer. Distribution of radial views and corresponding 2D real-time short axis images of closed-loop sampling at **A)** end-systole and **B)** end-diastole. Adaptive sampling results in near uniform radial distribution of views and thus high image quality. **C)** Cardiac motion is shown via projection through the left ventricle.

Despite being initialized with a golden angle trajectory, the closed-loop approach did not sustain this pattern and within a second the angular gaps deviated substantially from the golden trajectory. Reconstructed, multi-shot images illustrate that ECG cross-correlation can be used to robustly identify the correct cardiac phases. A projection through the left and right ventricle (along the 4-chamber view) is included to demonstrate the motion of the heart (**Figure 2C**). In this view, the motion of the ventricular wall is shown during the scan. In this healthy subject with no history of cardiovascular disease, the wall motion was normal with regular contractile function of the myocardium.

### Point spread function and View Distribution Analysis

During autonomous scanning the closed-loop controller selected new k-space radial projections on-the-fly. Since the user has no control over the trajectory, it was unclear what angles would be chosen and how it would affect image quality, uniformity and the point spread function. **Figure 3** shows that the autonomous scan achieved lower blurring (improved PSF images) than a similar segmented golden angle trajectory.

**Fig 3:**
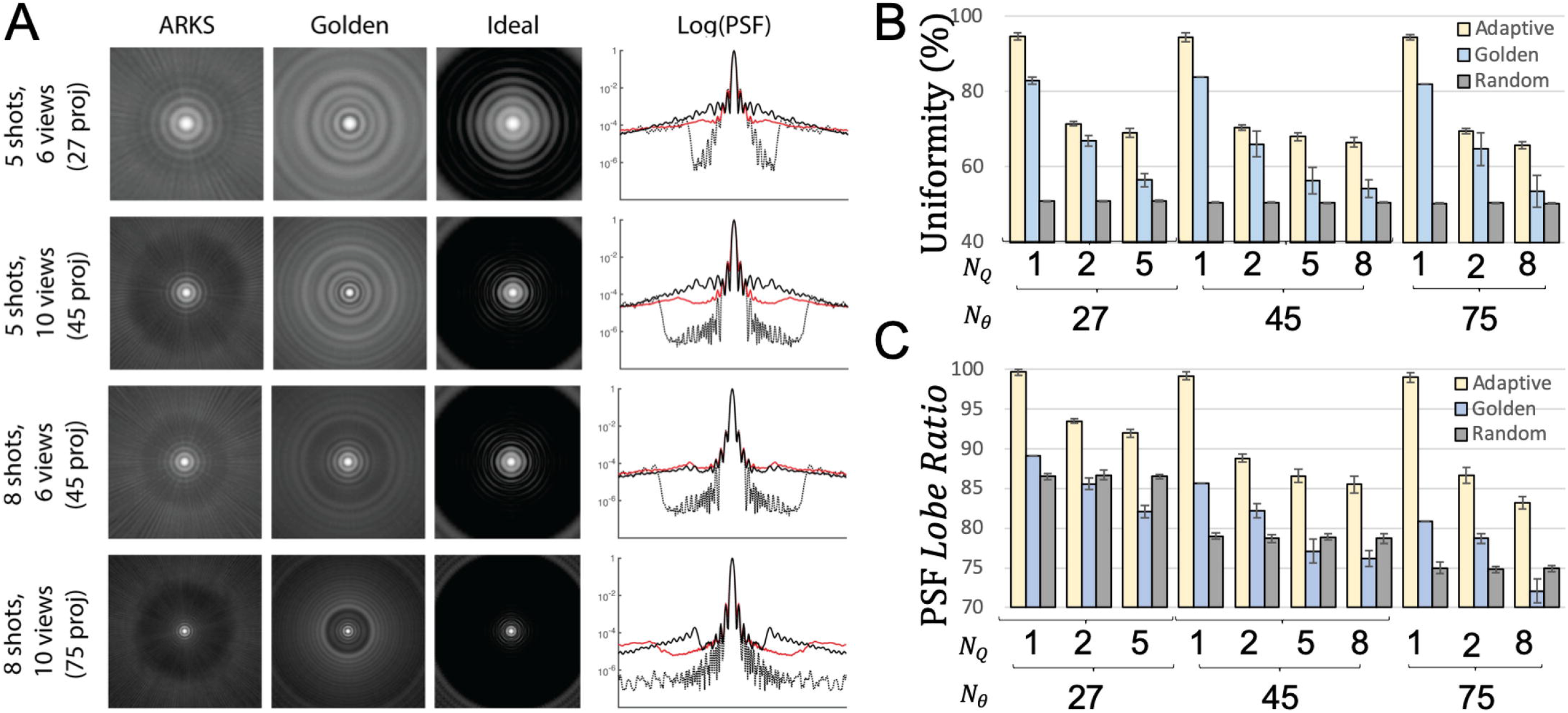
Point spread function and k-space sampling uniformity of autonomous, golden angle, random, and equispaced radial scanning. **Panel A)** Each row on the left shows the 2D point spread function for different segmentation strategies and each column shows autonomous (ARKS), golden or equispaced (Ideal) point spread functions. The fourth column shows a 1D profile for the circularly symmetric PSFs (autonomous shown in red). **Panel B and C)** K-space uniformity (Eq. 13) and the PSF (Eq. 9) for four subjects with different combinations of shots and segments. The autonomous approach results in improved PSF images (left), signal uniformity (top right), and point spread function lobe ratio (bottom right) across all combinations of shots and segments. ARKS = autonomous radial k-space sampling, PSF = point spread function, N_θ_ = number of radial views, N_q_ = number of shots

To obtain a better understanding of these properties, the uniformity of autonomous, golden and random trajectories was compared to the percent ideal uniformity that would be achieved with a constant Δ*θ*. If the trajectory were perfectly uniform, then all views would be equally distributed between 0 and 180°.

**Figure 3B** and **Tables 1** and **2** show that the autonomous scan had the best k-space sampling uniformity for all combinations of segmented trajectories that were investigated for both recorded ECGs (**Table 1**) and in-vivo testing (**Table 2**). Angular uniformity from autonomous sampling during recorded ECGs was 13±7% (range: 4 – 23%) higher than golden-angle sampling and PSF signal ratio was 14±6% (range: 8 – 26%) higher than golden-angle sampling. Single-shot closed-loop trajectories were only slightly lower in uniformity (94.4 – 94.6) and PSF signal (99.0 – 99.7) than equispaced sampling where the oldest view would be replaced immediately and surpassed golden angle trajectories in both metrics for all shot/segment combinations. In the four patients imaged in vivo (**Table 2)**, the autonomous approach increased k-space sampling mean uniformity 5±12% (range: -9 – 18%), decreased uniformity variability 45±14% (range: 32 – 58%), and increased PSF signal ratio 5±5% (range: 2 – 10%) relative to golden angle sampling.

### Arrhythmia Subject

To demonstrate the potential utility of this approach in imaging the heart of patients who have an arrhythmia, we utilized previously collected ECG and golden angle radial data (**Figure 4**) to demonstrate the closed-loop approach in the setting of a complex ECG. The cross-correlation algorithm (**Figure 4A**) correctly identified the correct phase in the cardiac cycle despite the complex arrhythmia seen on the ECG. While the data was acquired with golden-angle views, we calculated the angles the closed-loop approach would prescribe and show the short-axis (**Figure 4B**) and temporal projection images (**Figure 4C**). This suggests our approach could be used to increase the image quality of real-time images in patients with complex rhythms.

**Fig 4:**
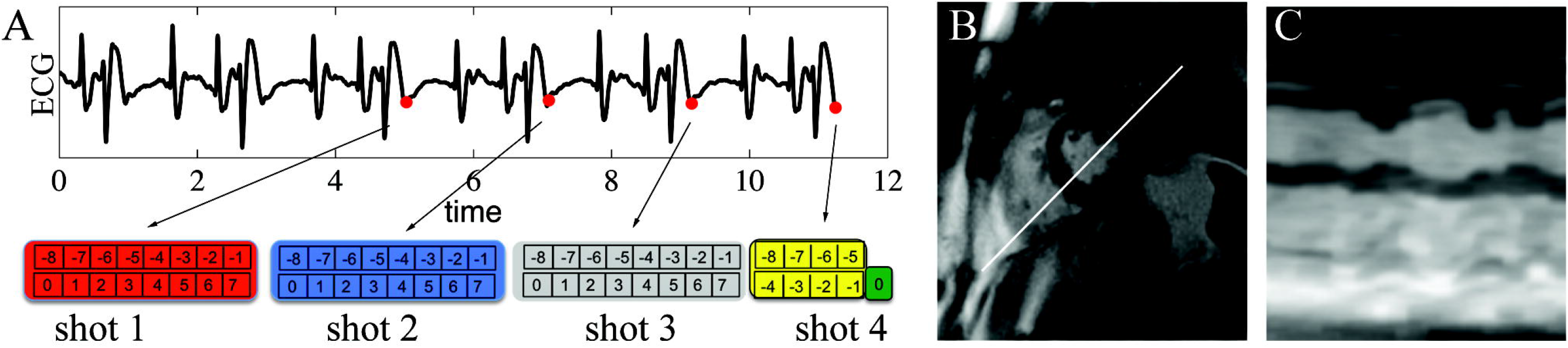
Utility of the proposed approach when imaging a patient with arrhythmia **A)** The cross-correlation based approach robustly identifies similar periods in the cardiac cycle in the setting of arrhythmia. **B)** A high-quality short axis image can be generated with the closed-loop scheme. **C)** The temporal projection illustrates the effect of complex rhythm on wall motion. This suggests the close-loop approach could enable multi-shot imaging of patients with complex rhythms.

## Discussion

We developed and investigated an autonomous k-space trajectory control system for cardiac MRI that implements closed-loop feedback. There were several important conceptual and technical aspects to this investigation. To our knowledge, this was the first implementation of an autonomous k-space trajectory using ECG and k-space trajectory feedback integrated in a closed-loop for segmented cine MRI. The algorithm was successfully implemented on a clinical whole-body MRI scanner as embedded and real-time software and its feasibility was shown in segmented cardiac cine MRI in healthy normal subjects. Our results showed that it enabled real-time cardiac MRI with good spatial and temporal resolution and reduced radial undersampling artifacts compared to conventional open-loop radial acquisitions.

The closed-loop radial trajectory is different from other radial sampling trajectories in that the view angle are not predetermined. For example, in an acquisition with fixed angular spacing Δ*θ*, the view angles increase from 0° to 180°- Δ*θ*. In a segmented acquisition with ten segments per heartbeat, the angles 0, Δ*θ*, 2Δ*θ*, …, 10Δ*θ* will be repeated for each frame of the first heartbeat and 11Δ*θ*, 12Δ*θ*, …, 20Δ*θ* for the second heartbeat, and so on, until all radial views are acquired. The total number of radial views *N* = 180/Δ*θ*, where both the angular spacing Δ*θ* and number of view angles *N* is determined before the scan begins. While this approach leads to the optimal sampling of k-space, the performance is substantially degraded if 1) one or more segments are not acquired due to arrhythmia rejection or 2) if significant motion – such as breathing – occurs due to arrythmia rejection causing a prolonged acquisition.

Similarly, in a golden angle trajectory [13], there is a fixed angular spacing Δ*θ* = 111.25° and each new angle is set by a schedule 0, Δ*θ*, 2Δ*θ*, …, and so on, before the scan begins. In the closed-loop trajectory, these view angles were not known in advance, but calculated on-the-fly using previous physiologic and k-space data. A limitation of predefined trajectories was that real-time physiologic information from the patient or knowledge about what and when data was collected was not used to judiciously inform the acquisition of new data. As was shown in the results, this led to sub-optimal k-space sampling behavior. In particular, golden radial sampling trajectory showed degraded image quality due to sub-optimal uniformity and high variability in sampling across cardiac periods. We showed that closed-loop sampling overcame this issue and achieved good uniformity and low variability in k-space sampling, permitting the synthesis of data across multiple cardiac periods for real-time acquisition and display with high image quality and low temporal footprint.

We observed some important differences between conventional segmented k-space sampling strategies and one that used a closed-loop. In segmented cine MRI, k-space data across several heartbeats would be combined to make a single dataset showing a single heartbeat. However, our approach has elements of both segmented and real-time acquisitions. Each image represented data from the previous 4 heartbeats, similar to how cine MRI would combine data from heartbeats, but the autonomous scan also results in images from every heart beat such as in a real-time acquisition. Furthermore, segmented cine MRI does not include closed-loop feedback systems for cardiac physiologic feedback such as from the ECG or from cardiac navigator signals. While prospective cine MRI certainly uses ECG feedback to provide synchronization for gating, it is open-loop since Cartesian spin warp imaging marches through a predefined list of phase encoding gradients [4,6] and non-Cartesian spatial encoding is performed using a similarly predefined list of view angles or spirals. In retrospectively gated reconstructions [6], ECG and navigator signals are used to properly bin acquired data into the correct cardiac or respirator phase, but they do not direct the sequence to update its trajectory in response to new information. Furthermore, both prospective and retrospectively gated cine MRI do not measure the output sampling trajectory nor do they maintain a desired setpoint for maintaining uniformity of k-space sampling across multiple heartbeats.

In real-time, interactive MRI, data is sampled as quickly as possible and images are displayed as soon as sufficient data has been collected. However, in many clinical applications, it is challenging to sample real-time MRI data with sufficient signal-to-noise ratio and spatial resolution without compromising temporal resolution. Furthermore, while recent advances in parallel and sparse scan acceleration techniques enable the collection of 2D images in real-time with good spatial and temporal resolution [1,27], image fidelity is degraded in real-time 3D applications. This framework could be adapted to improve both of these clinical applications.

This approach builds on features of other MRI techniques such automatic scanning [28,29] and view planning [28,30–32], inadvertent patient motion correction [5], and respiratory motion informed k-space sampling [33,34]. Furthermore, some acquisitions leverage use internal sensors to measure physiologic motion from k-space, image space navigators [5,35–38] or self-navigation [39] while other techniques use external sensors to capture motion information such as radiofrequency coils, ultrasound devices [40] and optical tracking devices [41,42]. Future work could integrate these physiologic signals into the closed-loop approach we describe.

While our results demonstrated the feasibility of an adaptive real-time system for cardiovascular MRI, additional work is needed to bring this technology into clinical use. Data should be gathered from patients to confirm that the real-time system provides accurate and reproducible beat-to-beat assessment of left ventricular function. Comparison of the sequence in patients with reduced ejection fraction or with left ventricular dyssynchrony should be performed to verify that temporal fidelity is sufficient to characterize compromised function or ventricular wall motion abnormalities. Further optimization of radiofrequency and gradient pulse durations is necessary to further reduce the sequence temporal footprint. The system should be integrated with a balanced steady-state free-precession pulse sequence because of its superior contrast, signal-to-noise ratio, and temporal resolution compared to spoiled gradient echo pulse sequences at 1.5 T. The cross-correlation algorithm appears to work well for subjects in sinus rhythms, however the length of the historical ECG data should be optimized for patients with arrhythmias or for 3D and interventional applications.

We implemented our technique in the setting of 2D radial k-space sampling for cine imaging since there is a straightforward definition of the optimal angle for subsequent acquisition. However, Cartesian and 3D sampling trajectories could both benefit from this approach. Furthermore, parametric mapping acquisitions such as T1-, T2-mapping and perfusion imaging are applications we plan to explore in future work. Lastly, this approach optimized sampling given past data acquisition without the expectation of future data acquisition. Other applications may allow for this assumption to be relaxed by modeling future data acquisition and further improve performance.

## Conclusions

We present an initial implementation of a closed-loop controller that defines radial k-space sampling. Based on recordings of ECGs in the MRI as well as 4 in vivo scans, the approach enables segmented acquisitions with improved sampling uniformity relative to the retrospective sorting of golden angle data. Furthermore, initial findings for a patient with arrhythmias suggest the approach would enable scanning of complex rhythms.

## Data Availability

Data cannot be shared publicly because of commercial research agreements with imaging vendors. Data are available (contact via Dr Contijoch) for researchers who meet the criteria for access
to confidential data.

## List of Abbreviations

DSP: digital signal process
ECG: electrocardiogram
FOV: field-of-view
RF: radiofrequency
PSF: point spread function
TE: echo time
TR: repetition time

## Declarations

### Ethics approval and consent to participate

In this study, all subjects and patients gave informed consent prior to participating in the study, approved by the Institutional Review Board of the University of Pennsylvania.

### Consent for publication

Not applicable

### Availability of data and materials

The datasets used during the current study are available from the corresponding author on reasonable request

### Author contributions

FC, YH, SKI, PK, GG, MAE, SB, and WRW contributed to the conception and design of the work, acquisition and analysis of the data, interpretation of the data, and drafting/revising of the manuscript. JHG, RCG, JJP contributed to the conception and design of the work, interpretation of the data, and drafting/revising of the manuscript. All authors approve the submitted version and ensure the integrity of the work.

## Acknowledgements

Not applicable

## Figure Captions

**Supp Fig 1:** Closed-loop identification of prior cardiac phases **A**, (top) A brief portion of ECG signal is compared to the entire scan ECG signal (bottom) via cross correlation. **B**, (top) Cross-correlation output. Local peaks are indicated in red. (bottom) In this segmented acquisition, MRI data from 4 beats was used for reconstruction. Radial views from shots 1 through 3 (red, blue and gray) are used to determine the newest segment (green). The number of radial views obtained from each shot (e.g. 8 views/shot) can be varied depending on the application.

**Supp Fig 2:** Training mode A consists of TCP/IP reading of new ECG data, buffer storage of ECG sampled, and other software overhead. Training mode B occurs once the buffers are populated and includes calculation of the cross correlation to identify similar periods of ECG signal. During training mode B, the optimal angle is not calculated since insufficient beats are identified. Active mode occurs after both the buffers are populated and sufficient number of beats can be identified. It includes calculation of the optimal sampling angle.

**Supp Fig 3:** Radial k-space sampling uniformity analysis. **A**, histogram of view angle gaps Δ*θ* for uniform (black) and random (red) radial sampling trajectories. The uniform sampling distribution is a delta function positioned at the 180°/N_*θ*_. **B**, cumulative distribution functions (CDF) for uniform (black) and random (red) radial sampling trajectories. The uniformity *U* = *CDF*(*χ*_*e*_)/*CDF* (*χ*_0_).

**Supp Movie 1:** ECG sampling, results of the cross-correlation, k-space sampling, and imaging results over 10 seconds for the patient shown in Figure 2.

